# Immune alterations during SARS-CoV-2-related acute respiratory distress syndrome

**DOI:** 10.1101/2020.05.01.20087239

**Authors:** Lila Bouadma, Aurélie Wiedemann, Juliette Patrier, Mathieu Surénaud, Paul-Henri Wicky, Emile Foucat, Jean-Luc Diehl, Boris P. Hejblum, Fabrice Sinnah, Etienne de Montmollin, Christine Lacabaratz, Rodolphe Thiébaut, JF Timsit, Yves Lévy

**Affiliations:** APHP- Hôpital Bichat – Médecine Intensive et Réanimation des Maladies Infectieuses, Paris, France; UMR 1137 - IAME Team 5 – DeSCID: Decision Sciences in Infectious Diseases, Control and Care, Inserm/Univ Paris Diderot, Sorbonne Paris Cité, Paris, France; Vaccine Research Institute, Université Paris-Est Créteil, Faculté de Médecine, INSERM U955, Créteil, France; APHP, Hôpital Georges Pompidou, Médecine Intensive Reanimation, Paris France; Univ. Bordeaux, Department of Public Health, Inserm Bordeaux Population Health Research Centre, Inria SISTM, UMR 1219; Vaccine Research Institute (VRI), Créteil, France; CHU Bordeaux, Bordeaux, France; Assistance Publique-Hôpitaux de Paris, Groupe Henri-Mondor Albert-Chenevier, Service Immunologie Clinique, Créteil, France

## Abstract

We report a longitudinal analysis of the immune response associated with a fatal case of COVID-19. This patient exhibited a rapid evolution towards multiorgan failure. SARS-CoV-2 was detected in multiple nasopharyngeal, blood, and pleural samples, despite antiviral and immunomodulator treatment. Clinical evolution in the blood was marked by an increase (2-3 fold) in differentiated effector T cells expressing exhaustion (PD-1) and senescence (CD57) markers, an expansion of antibody-secreting cells, a 15-fold increase in γδ T-cell and proliferating NK-cell populations, and the total disappearance of monocytes, suggesting lung trafficking. In the serum, waves of a proinflammatory cytokine storm, Th1 and Th2 activation, and markers of T-cell exhaustion, apoptosis, cell cytotoxicity, and endothelial activation were observed until the fatal outcome. This case underscores the need for well-designed studies to investigate complementary approaches to control viral replication, the source of the hyperinflammatory status, and immunomodulation to target the pathophysiological response.

## Introduction

First reported in December 2019 in China^1,2^, SARS-CoV-2 (a beta coronavirus) can cause a respiratory syndrome that manifests a clinical pathology resembling mild upper respiratory-tract disease (common cold-like symptoms) and occasionally severe lower respiratory-tract illness and extra-pulmonary manifestations, leading to multiorgan failure and death. Such a severe clinical condition displayed by certain patients affected with COVID-19 pneumonia are strongly reminiscent of previous and recent epidemic cases of respiratory failure associated with related coronaviruses, such as MERS-CoV and SARS-CoV^3^

In more severe cases, infection can cause pneumonia, severe acute respiratory syndrome, kidney failure, and even death^3^. Death results from hypoxemic respiratory failure in patients developing severe acute respiratory distress syndrome (ARDS) and is associated, in a substantial portion of patients, with an inflammatory syndrome and cytokine storm^4^ that may originate from immune cells^5^. Severe and critical COVID-19 pneumonia shares features with “cytokine storms”, such as those seen in severe cytokine-release syndrome, which is characterized by fever, hypotension, and respiratory insufficiency associated with elevated serum cytokine levels, including those of IL-1 and IL-6, mostly produced by myeloid cells^6^. Previous studies from SARS animal models and infected humans have suggested the occurrence of an aberrant host cytokine storm, resulting in an excessive clinical manifestation that plays a critical role in disease severity^7–9^. However, a precise description of the immune mechanisms responsible for the pathophysiology and acute mortality in COVID-19-infected patients is still unavailable. Here, we provide a full description of a fatal case of COVID-19, including a chronological immune profile of the patient, to aid the determination of the putative underlying mechanisms of this deadly case and the identification of potential therapeutic targets.

## Results

### Patient

An 80-year-old male visited an emergency hospital department in early 2020. His symptoms had commenced three days earlier with fever and diarrhea (day 0) (Fig. 1). At entry, he presented purulent sputum and dyspnea. He did not report any specific exposure within the 14 days prior to the onset of symptoms. Clinical examination revealed a temperature of 37.2°C, oxygen saturation of 88% while breathing ambient air, pulse of 65 beats/minute, blood pressure of 132/82 mmHg, and respiratory rate of 17 breaths/minute. A medical examination suggested a lower respiratory-tract infection. Routine blood tests showed severe hypoxemia, with a PaO_2_ of 53 mmHg in ambient air, requiring oxygen therapy with a nasal cannula (4 L/min), and an elevated level of C-reactive protein of 124 mg/L. Kidney function and hepatic tests were in the normal range. The general biological data were previously reported^10^. Chest radiography showed bilateral alveolar opacities.

**Fig. 1.**
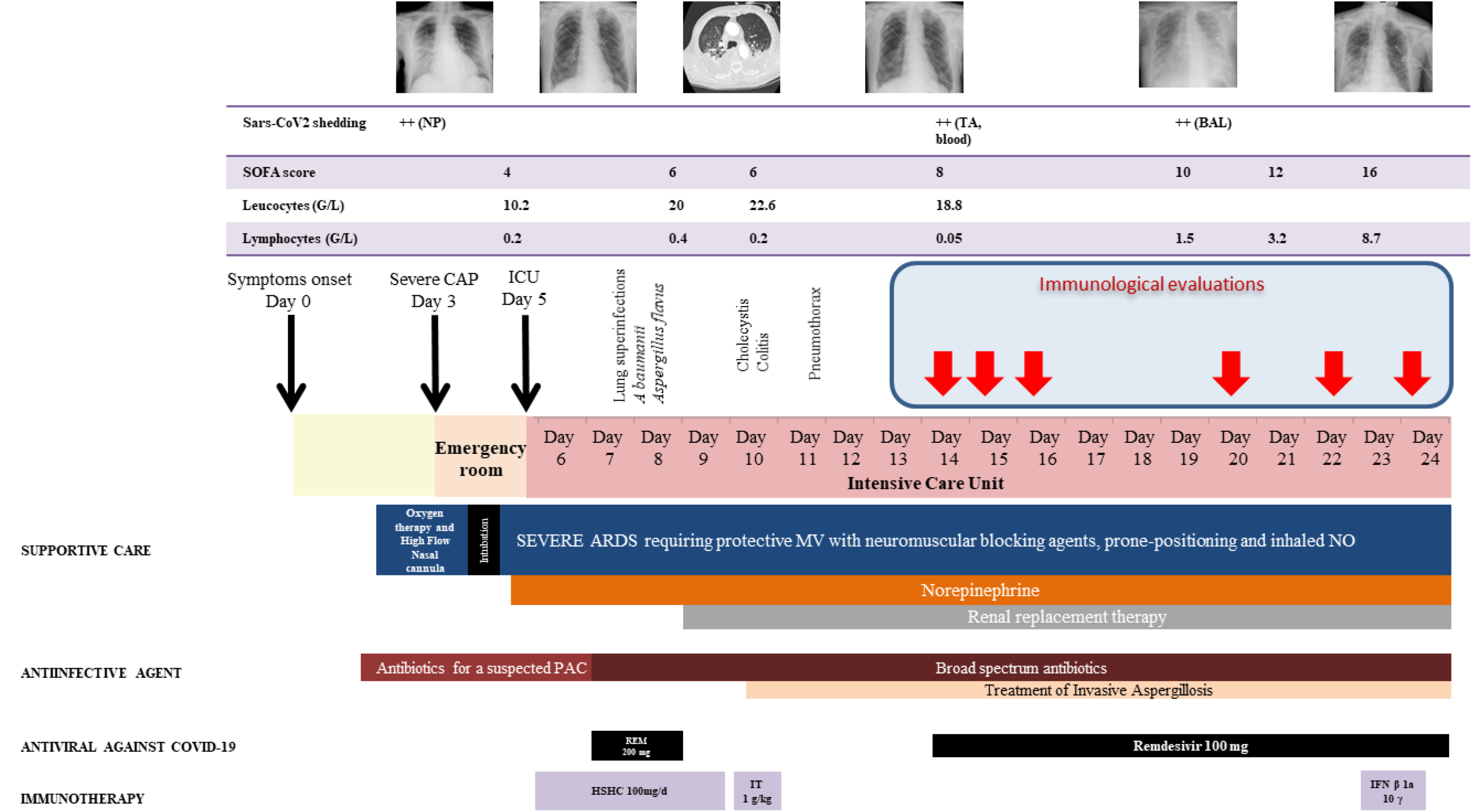
Timeline from arrival in Europe to death in the ICU. TA: tracheal aspirate, SOFA: sepsis-related organ failure assessment, CAP: community acquired-pneumonia, MV: mechanical ventilation, NO: nitric oxide, REM: remdesivir, HSCH: hydrocortisone hemisuccinate, IT: immunoglobulin therapy, IFN β1a: interferon β1a

As he did not fulfill the case definition, his case was not immediately considered to be one of possible COVID-19 infection. Nonetheless, precautions to avoid airborne and contact contamination were observed while awaiting a COVID-19 test result. Community-acquired pneumonia was diagnosed and treated with amoxicillin-clavulanate. Classic etiological agents were ruled out by real-time multiplex PCR screening. On day 5, he developed a fever and acute respiratory failure and was transferred to the ICU. He subsequently developed multiorgan failure with ARDS, acute kidney injury, liver failure, and sepsis-like shock and was consequently placed under protective mechanical ventilation and vasopressors on day 6 (Fig. 1). The COVID-19 diagnosis was confirmed on day 7. Broad-spectrum antibacterial and Remdesivir were started and then adapted following identification of susceptible *Acinetobacter baumannii* (Filmarray multiplex PCR confirmed by tracheal aspirate culture) and *Aspergillus flavus* (tracheal aspirate culture). On day 9, Remdesvir was stopped and the patient required renal replacement therapy. A CT-scan on day 10 showed bilateral pleuro-pneumopathy, associating pleural effusion, alveolar condensations, ground glass opacity, and pulmonary cysts. Colitis and cholecystitis were also diagnosed, leading to broadening of the antimicrobial treatment.

SARS-CoV-2 shedding was detected throughout the follow-up in all daily nasopharyngeal swabs collected from day 7 to the patient’s death and also several times in blood and pleural effusion samples but not in urine or rectal swabs. On day 14, Remdesivir was re-initiated. Although we found no other superinfection, despite multiple investigations (Fig. 1), the patient’s condition worsened. One dose (10 µg) of interferon β-1a was administered on day 23. The patient died on day 24 from massive hemoptysis and uncontrolled multiorgan failure.

### Kinetics of immune and inflammatory responses

We longitudinally analyzed the blood immune response during worsening of the clinical status and progression of the COVID-19 infection. The frequency of naïve CD4^+^ and CD8^+^ (CD45RA^+^CCR7^+^) T cells was low on day 14 (24.2% and 6%, respectively) relative to that of five healthy donors (HD) (Median [Interquartile Range (IQR)] (42.9% [35.3-49.8] and 29.5% [13.4-45], respectively) and remained so to the end of the follow-up. In contrast, the frequency of CD4^+^ and CD8^+^ (CCR7^-^CD45RA^-^) effector memory T cells was high on day 14 (51.7 and 28.6%, respectively) relative to that of HD (28.2% [21.8-29.4] and 23.5% [11-26], for CD4^+^ and CD8^+^ T cells, respectively) and peaked at day 24 (67.4% and 46.2%, respectively) (Fig. 2A).

**Fig. 2.**
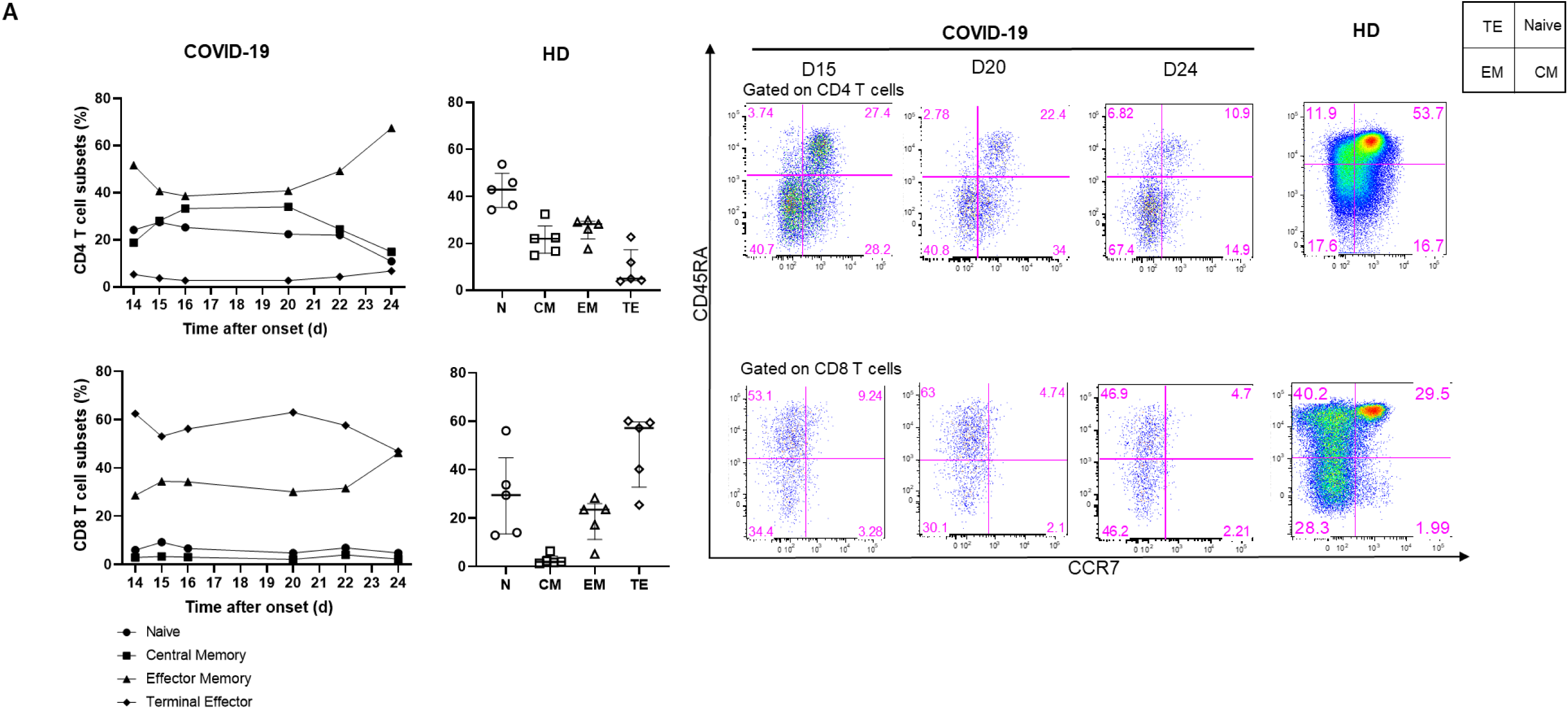
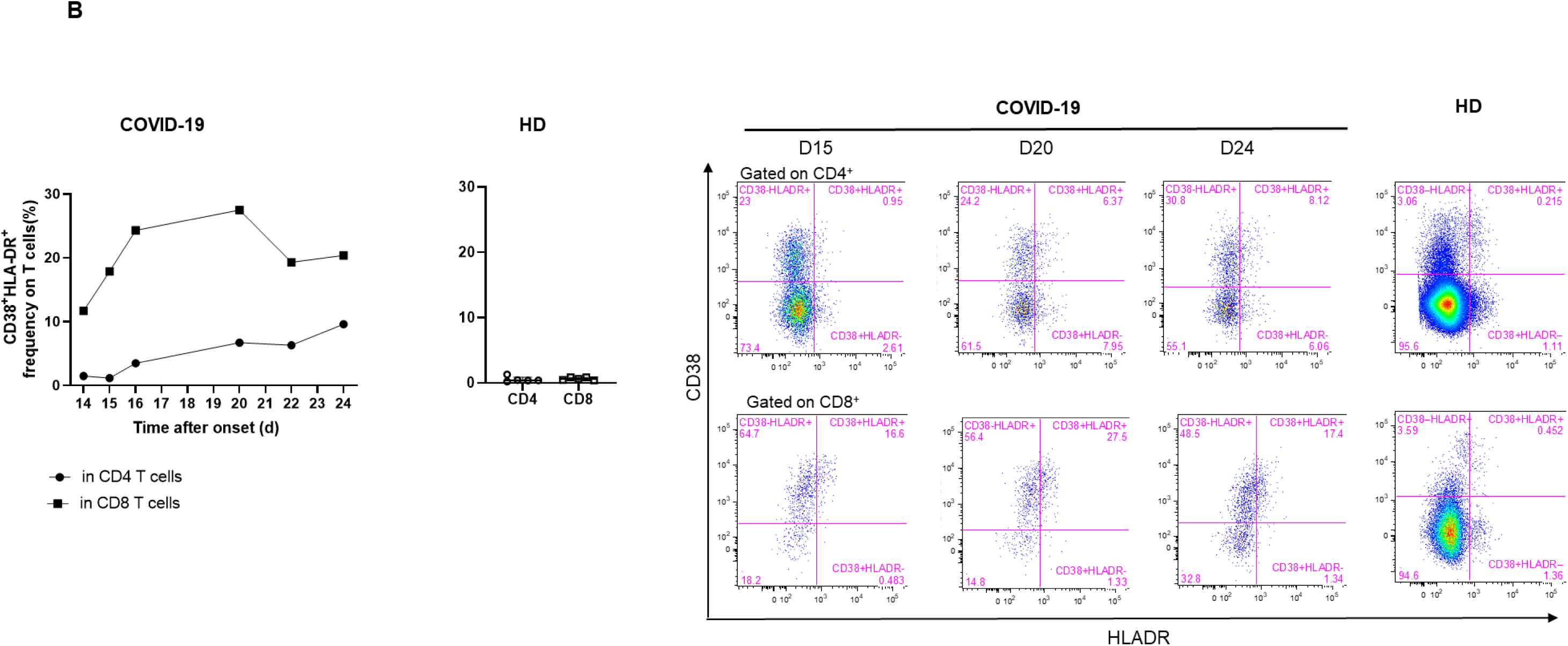
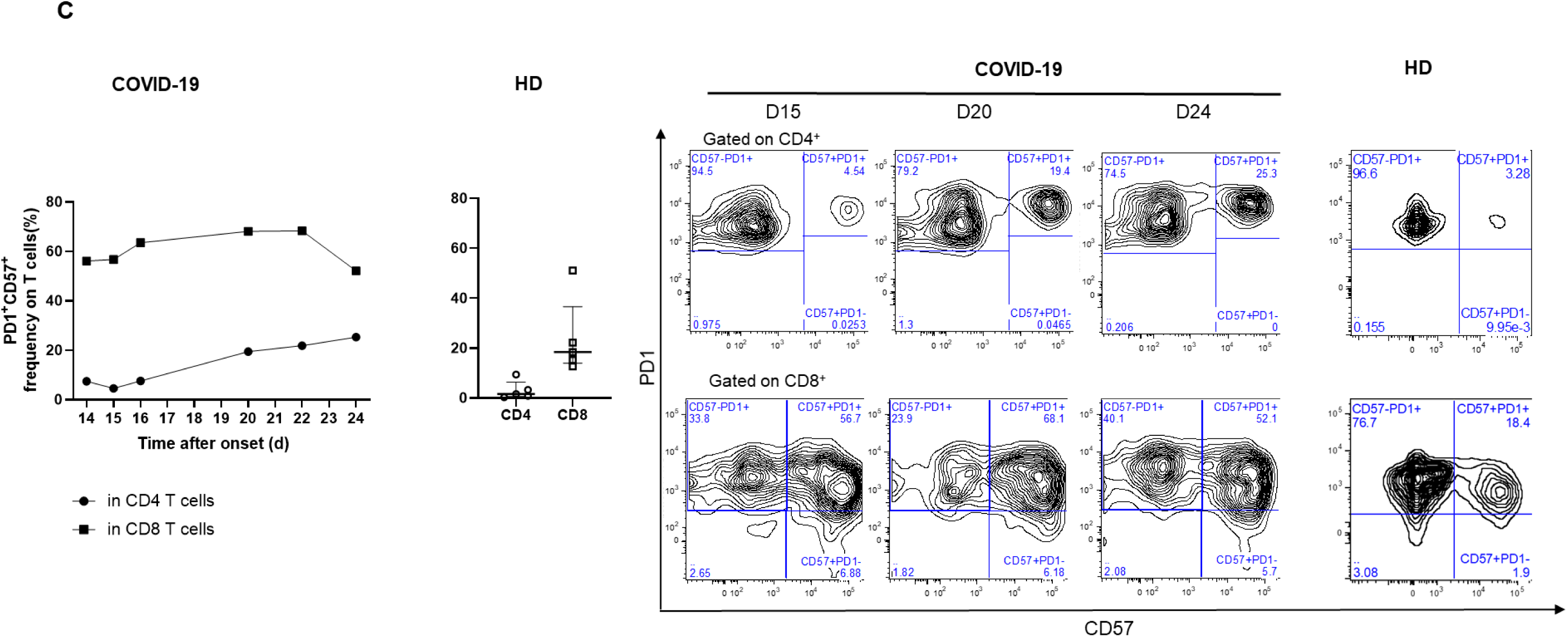
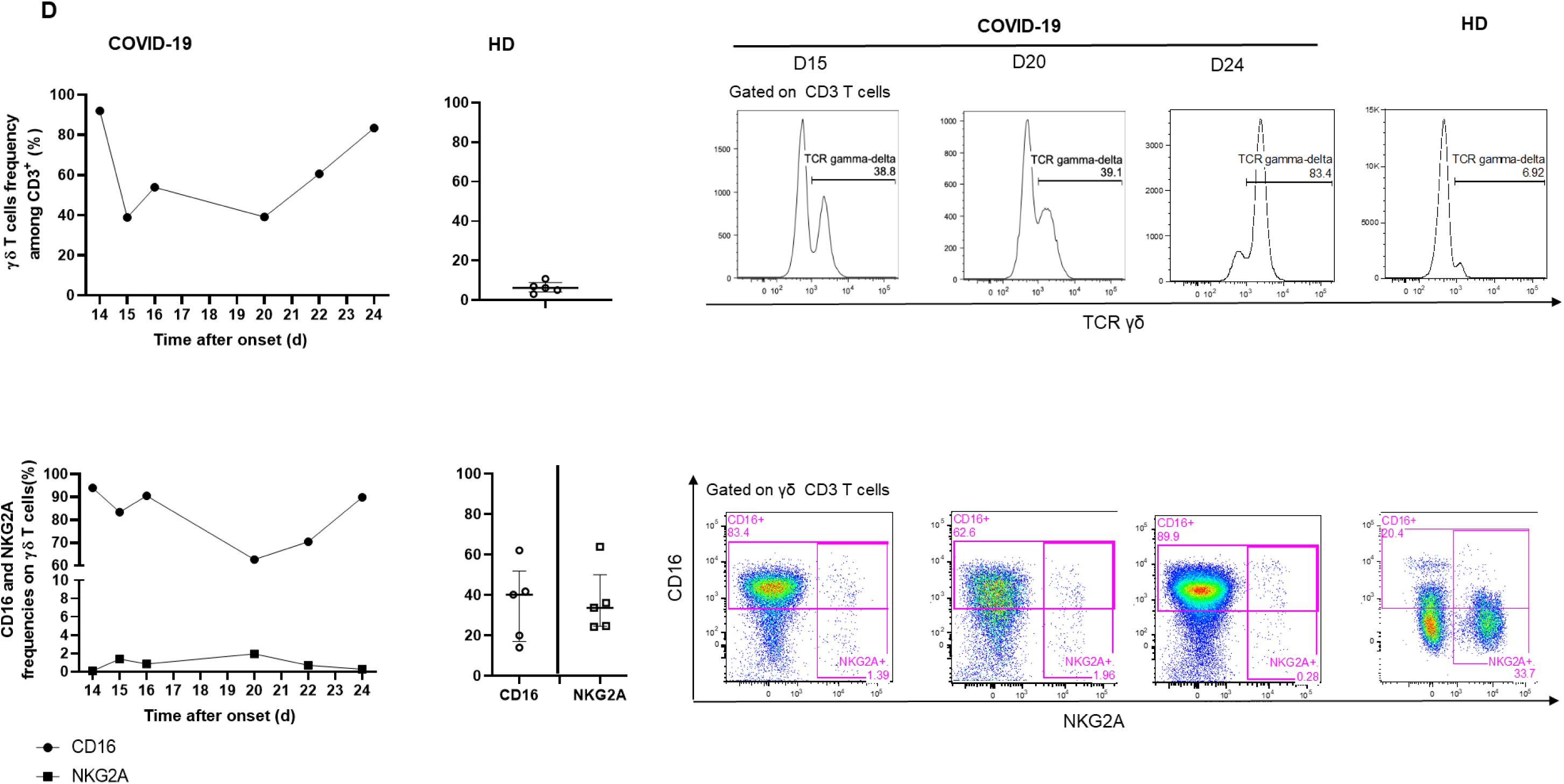
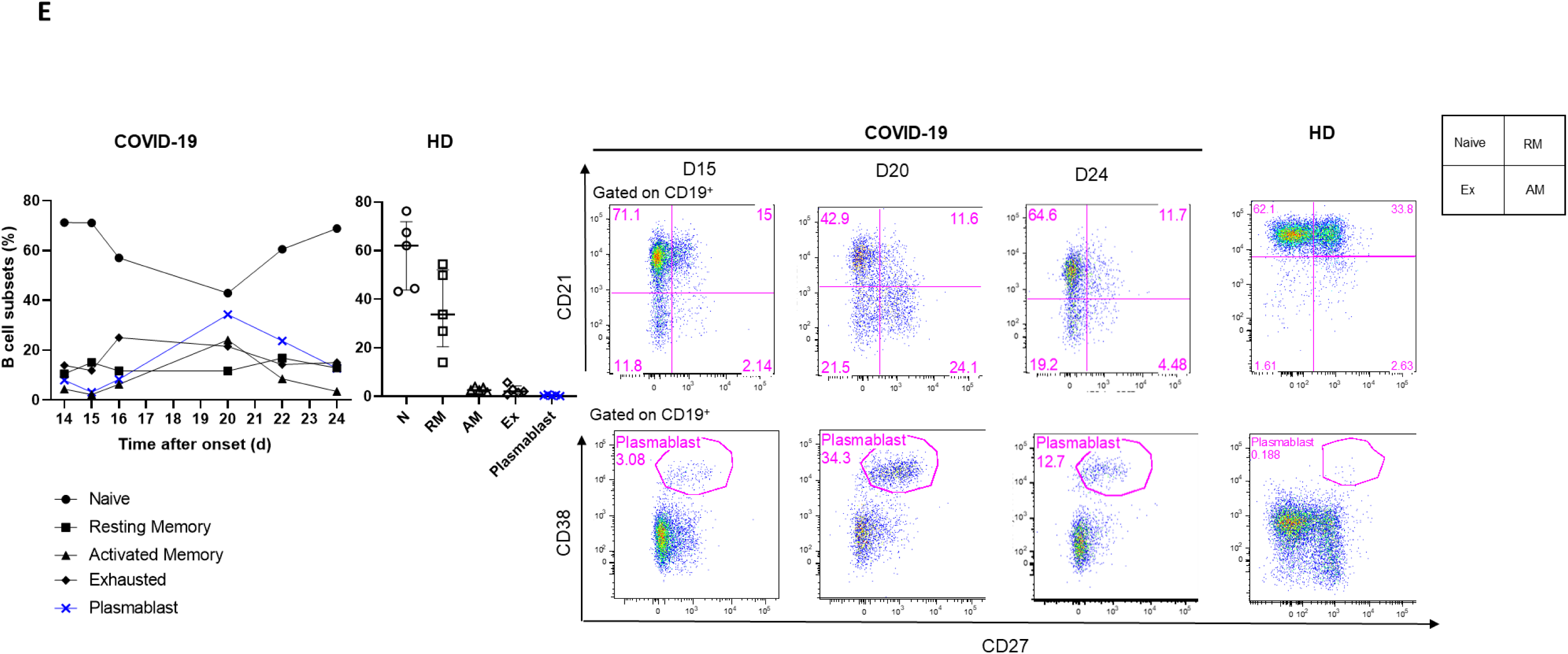
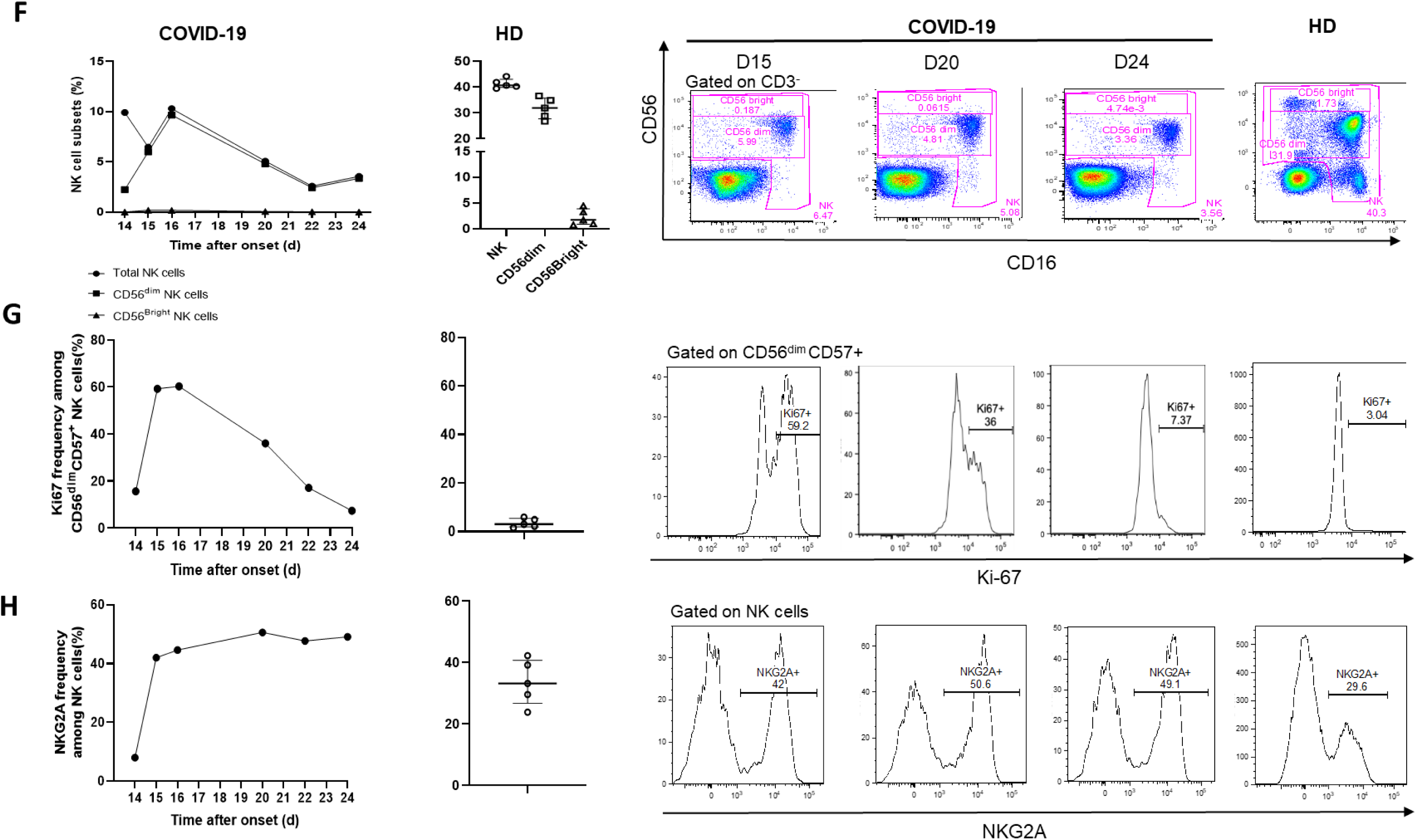
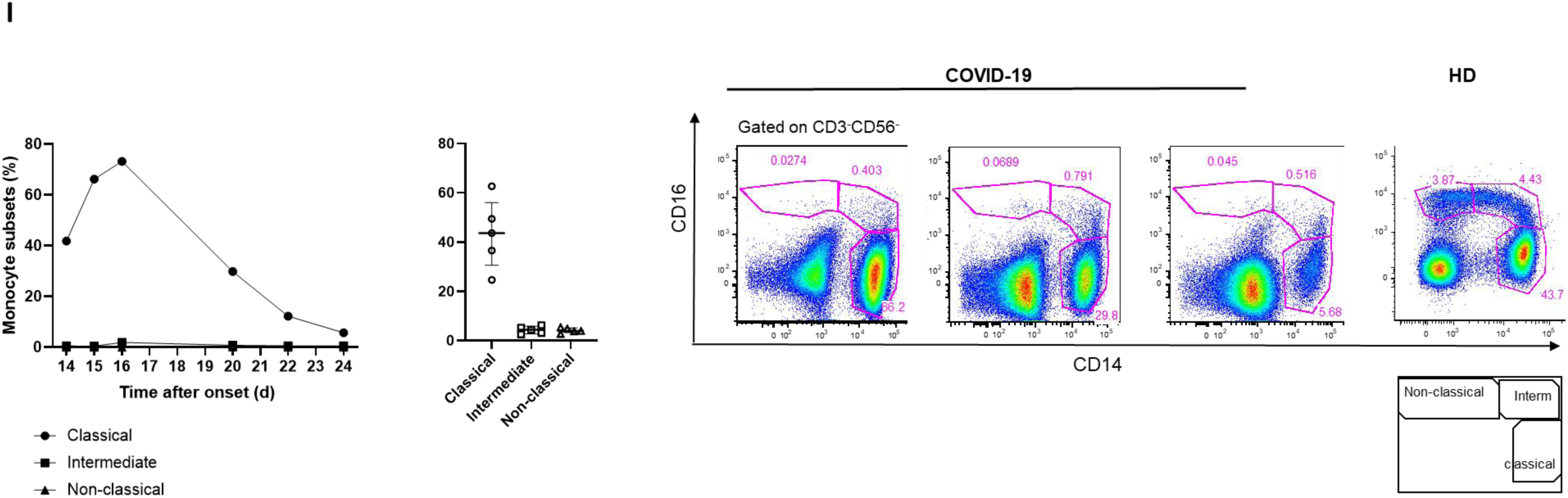
Kinetics and activation status of immune-cell subsets throughout the infection. **A-C** Frequency (left set of plots) of CD4 and CD8 T-cell subsets (A), activated CD38^+^HLADR^+^ (B), and exhausted PD1^+^CD57^+^ CD4 and CD8 T cells (C). **D** Frequency of γδ T cells (gated on CD3^+^ T cells) and CD16 and NKG2A expression (gated on γδ CD3 T cells). **E** Frequency of B-cell subsets and plasmablasts (CD38^++^CD27^+^) gated on CD19^+^ B cells. **F-H** Frequency of NK-cell subsets (gated on CD3^-^) (F), differentiated Ki67^+^ NK cells (gated on CD56^dim^CD57^+^ NK cells) (G) and inhibitor receptor NKG2A (gated on NK cells) (H). **I** Monocyte subsets (gated CD3^-^CD56^-^) detected by flow cytometry of blood collected at days 14-20 following symptom onset from the patient and healthy donors (n = 5, median with interquartile range); gating examples shown to the right.

The profile of the patient showed dramatically marked cell activation relative to HD, with an expansion of CD38^+^HLA-DR^+^ CD8^+^ T cells during the follow-up. Activated CD8^+^ T-cell levels peaked on day 20 (27.5% and 0.6% [0.4-0.8], for the COVID-19 patient and HD, respectively) (Fig. 2B). Similarly, the frequency of exhausted and senescent CD4^+^ and CD8^+^ T cells (PD-1^+^CD57^+^) increased in the COVID-19 patient from day 14 (7.43 and 56.1%, respectively) throughout the follow-up, peaking on day 24 for CD4^+^ T cells (25.3%) and day 22 for CD8^+^ T cells (68.3%) and were dramatically higher than in the HD (1.6% [0.6-6.3] and 18.4%, [14-36], respectively) (Fig. 2C).

We also explored the proportion of γδ T cells among CD3^+^ T cells. The frequency of γδ T cells was markedly high on day 14 (92%), relative to that of the HD (6.2% [4-8.9]), lower on day 15 (38.8%), and increased continuously from day 20 until day 24 (83.4%). The phenotype of the γδ T cells was characterized by high levels of the marker CD16 but not the inhibitory receptor NKG2A (83.4 and 1.4%, respectively) on day 15 relative to that of HD (40.2% [17-52] and 33.7% [24.5-50] for CD16 and NKG2A, respectively) which persisted until death (Fig. 2D).

The phenotype of the B cells evolved significantly. The frequency of activated memory (CD19^+^CD27^+^CD21^-^) B cells and plasmablasts /antibody-secreting cells (ASC) (CD19^+^CD38^+^CD27^+^) peaked on day 20 (24.1 and 34.3%, respectively) and was markedly higher than that in HD (2.6% [2.1-3.8] and 0.32% [0.25-0.75], respectively), as was that of exhausted CD19^+^CD27^-^CD21^low/neg^ B cells (21.5% versus 2.1% [1.2-4.4] in HD) (Fig. 2E).

The total NK cell frequency decreased from day 16 (10.3%) to day 24 (3.6%) and remained lower than in HD (40.7% [40-43]), with a much lower frequency of NK CD56^dim^ cells than in HD, with a nadir at day 14 (2.2%), associated with the total absence of NK CD56^bright^ cells (32% [28-36] and 1.7% [0.9-3.8] in HD, respectively) (Fig. 2F). Remarkably, a high frequency of differentiated CD56^dim^ CD57^+^ NK cells were cycling (Ki67^+^) on day 14 (15.6%), peaked on day 16 (60.3%), and then continuously decreased to day 24 (7.37%) (Fig. 2G). The frequency of NK cells expressing the inhibitory marker NKG2A markedly increased from day 14 (8%) to day 15 (42%), peaked on day 20 (50.6%), and remained much higher than in HD (33% [27-41]) (Fig. 2H).

Finally, an analysis of monocyte populations also showed dramatic changes throughout the follow-up. The frequency of classical CD14^+^CD16^-^ monocytes steadily decreased from day 16, through days 20 and 22, to day 24 (73.2, 29.8, 12.2, 5.7%, respectively), becoming much lower than in HD (43.7% [31-56]). Strikingly, the frequency of intermediate CD16^+^CD14^+^ and non-classical CD14^-^CD16^+^ monocytes remained extremely low throughout the follow-up, with a nadir at day 15 (0.4 and 0.02% versus 4.4% [2.9-5.8] and 4% [3.3-5.1] for HD, respectively) (Fig. 2I).

In addition, we longitudinally measured the levels of 72 analytes in the serum of the COVID-19 patient and compared them to those of HD (n = 5) and patients with septic shock (SS, n = 5). Most of these markers were found at much higher levels than in HD and/or SS patients (Fig. 3). Moreover, repeated measurements allowed us to study the kinetics of the various profiles. We detected a storm of pro-inflammatory and Th1/Th2 factors on day 14 after onset, of which some tended to decrease throughout the follow-up, while remaining higher than in the HD and/or patients with SS (IFN-γ, MIP-1α, MIP-1β, TGFα, MCP-1, TNF-α, IL-1α, β-NGF, Basic FGF, IFN-α2, IL-5, G-CSF), as well as a burst of Th1 cytokines (IL-2, IL-3, IL-12 (p70)), Th2 cytokines (IL-4, IL-5, IL-6), and an immune-modulator (IL-1RA). Some of these factors persisted at a lower level on day 15 post illness, but the profile was significantly enriched by an increase in the level of other markers (4-1BB/TNFRSF9/CD137, GM-CSF, Midkine, IL-21, Flt-3 Ligand, CCL28, Fas Ligand/TNFSF6, IL-17E/IL-25, IL-23, CD40 Ligand/TNFSF5, CXCL14/BRAK, IL-31, Granzyme A, PD-L1/B7-H1) associated with T cell activation, exhaustion, and apoptosis. Of note, IL-1RA decreased dramatically from day 15 to day 22, contrasting with persistently high levels of IL-1. On day 20, we observed a significant increase in the level of biomarkers of cell cytotoxicity, neutrophil chemotaxis, and endothelial activation (MIG, VEGF, IL-7, Granzyme B, GRO-a, PDGF-BB, RANTES, IL-8, IL-9, EGF). On day 24 (death), after one dose of IFN-β 1a on day 23, there was a dramatic increase in the level of several cytokines, reflecting the activation of T cells, monocytes, and inflammation (IL-2, IP-10, TRAIL, IL-17, IL-12(p70), CD163, IL-12 (p40), IL-15, TNF-β, SDF-1a, LIF, IL-1β), as well as an anti-inflammatory profile (IL-3, IL-4, IL-13, IL-1RA) and that of a leaky gut (I-FABP).

**Fig. 3.**
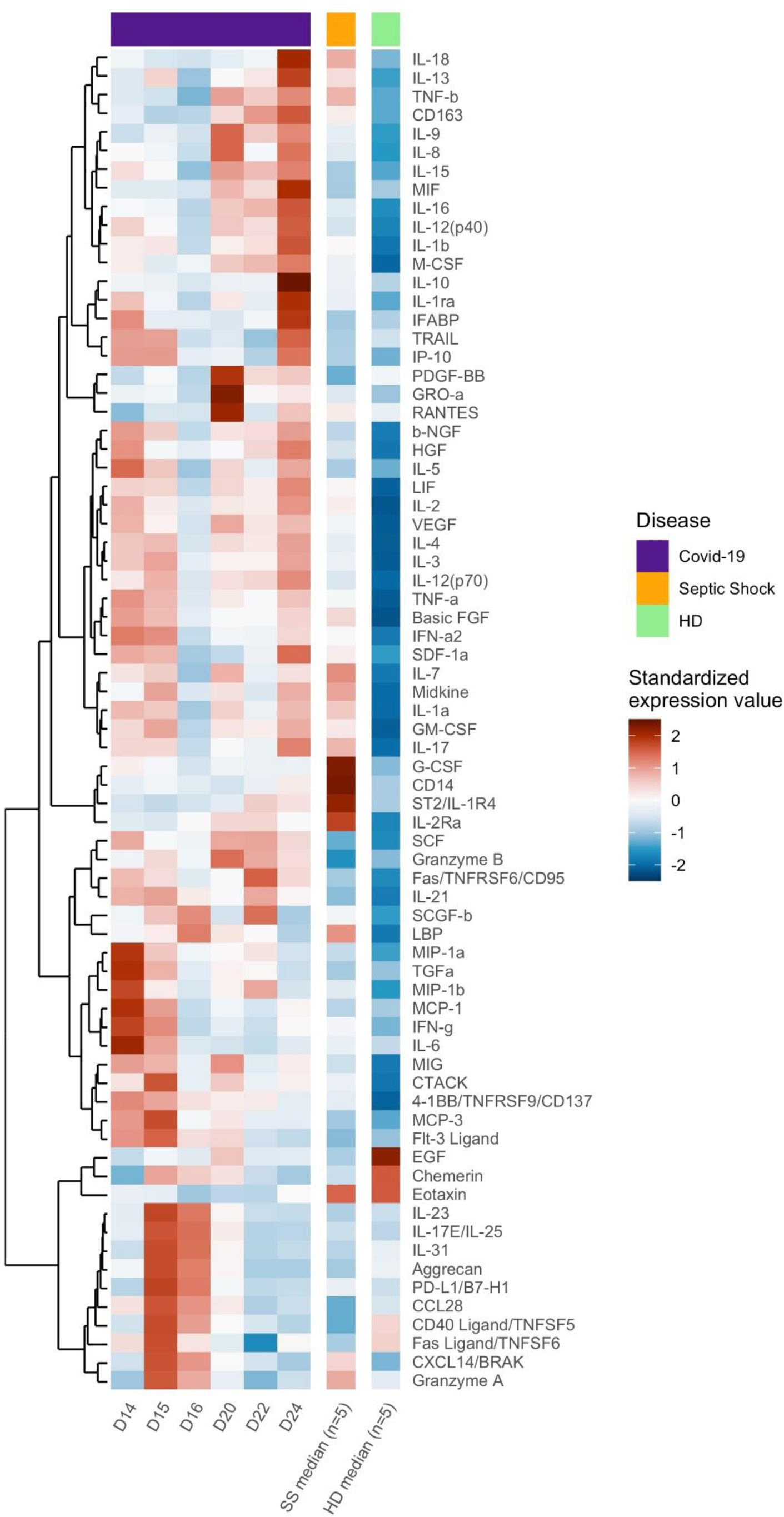
Heatmap of standardized biomarker expression in serum throughout the infection. The colors represent standardized expression values centered around the mean, with variance equal to 1. Biomarker hierarchical clustering was computed using the Euclidean distance and Ward’s method^31^. HD: Healthy donors (n = 5), SS: Septic Shock (n = 5)

## Discussion

This report provides a dynamic overview of immune abnormalities associated with the clinical outcome of a fatal case of COVID-2019. The lack of knowledge of the pathophysiology of this new infection, the severity of the cases, and the absence of confirmed therapies have been critical challenges for all physicians and intensivists since the beginning of this year. In an epidemic context, it is crucial to rapidly improve our understanding of the disease to better manage patients and to share knowledge on the most severe cases and learn from them.

From a clinical prospective, the presentation of a COVID-19 infection resembles a typical community-acquired severe respiratory infection that occurs one week after the onset of nonspecific flu-like symptoms, but exhibiting a wider range of severity than MERS-CoV and SARS-CoV-1 infections^11,12^. The reason for such marked heterogeneity in individual sensitivity to COVID-19 and the potential roles of ageing and comorbidities, beyond their association with clinical worsening, are poorly understood. The patient described in this report exhibited a rapid and fatal evolution towards multiorgan failure, with long and sustained persistence of SARS-CoV-2 nasopharyngeal shedding associated with viral RNA detection in multiple blood and pleural effusion samples. Although neither viral quantification nor the detection of infective virus were performed in this case, its persistence is consistent with a recent report suggesting that higher viral loads may be associated with severe clinical outcomes^13^.

As described for patients suffering from SARS, MERS, and COVID-19, the case presented here exhibited persistent lymphopenia. Although, this observation is commonly reported in several severe viral illness^14^, persistent lymphopenia three days after ICU admission is associated with an increased risk of ICU-acquired infection and is a predictive factor for increased 28-day mortality^5,15^. Thus, a lymphocyte count performed early after admission is a simple blood test that could be used as a prognostic marker to possibly explain early co-infections in severe viral infections. Moreover, a multicenter retrospective study showed that lower lymphocyte counts and underlying comorbidity, older age, and higher LDH levels at presentation are independent high-risk factors for COVID-19 progression. The authors developed the CALL score (comorbidity, age, lymphocyte count and LDH level), which ranges from 4 to 13 points, with a cutoff value of 6 points. The positive and negative predictive values were 50.7% (38.9%-62.4%) and 98.5% (94.7-99.8), respectively. Importantly, a lymphocyte count ≤ 1 10^9^/L is worth 3 points in the CALL score.

The data acquired during previous Coronavirus infections (SARS and MERS-CoV) did not allow any conclusions to be drawn concerning the pathophysiology of these infections. However, the balance between viral factors and the host response, when the latter is exacerbated, appears to be crucial in the evolution of the clinical course of the disease and prognosis. The intensity of the inflammatory response may play a role in the pathogenesis of SARS-CoV2, in particular in the airways. An in-depth longitudinal analysis of the immune profiles of our case showed dramatic alterations in the homeostasis of all blood cell populations, reflecting severe disturbances in innate and adaptive immunity. Some are highly compatible with an uncontrolled viral infection, as exemplified by an increase (2-3-fold) of most differentiated effector memory CD4^+^ and CD8^+^ T cells. Most of these cells expressed characteristics of exhausted (high level and frequency of PD-1) and senescent cells (CD57 marker).

Consistent with these observations and as previously reported in one case with moderate COVID-19 infection^16^, we also show an expansion of ASC and exhausted memory B cells. We did not assess the humoral response of our patient because a validated serological test was not yet available. A recent report has demonstrated the role of anti-spike IgG in severe acute lung injury by skewing the inflammation-resolving response by macrophages in SARS-CoV-2 macaque models^17^. In this study, the presence of anti-spike IgG prior to viral clearance abrogated the wound healing response and promoted MCP-1 and IL-8 production and pro-inflammatory monocyte/macrophage recruitment and accumulation. This phenomenon has also been suspected in deadly SARS-CoV infections. It would likely be informative to study the functional profile of specific IgG in COVID-19 infected patients with a rapid and severe clinical evolution.

Strikingly, we observed a dramatically high level of γδ T cells throughout the clinical course of the disease, up to 15-fold higher than in HD. γδ T cells were first described in the lung and shown to make up 8 to 20% of CD3^+^ cells^18^ and play critical roles in anti-viral immune responses, tissue healing, and epithelial cell maintenance^19^. This observation is compatible with previous reports in convalescent health workers infected with SARS-CoV-1, who exhibited an expansion of Vγ9Vδ2 T cells able to inhibit SARS-CoV replication and kill SARS-CoV–infected target cells^20^. In our patient, the γδ T cells expressed activation markers (CD16) but only low levels of the inhibitory receptor NKG2A, suggesting that they may have exhibited a killing capacity. Analyses of the T-cell repertoire and functional profile of these cells need to be performed in a larger cohort of patients.

Along with these T cell populations, NK cells and monocyte/macrophages in the lung are at the first line of defense against pathogens. These cells produce a large array of pro-inflammatory cytokines. The high level of NK-cell proliferation and their disappearance from the blood suggest trafficking to the lung, which may contribute to lung-cell cytotoxicity. Three populations of monocytes, CD14^+^CD16^-^, CD14^+^CD16^+^, and CD14^-^CD16^+^, were at low or undetectable levels, also suggesting trafficking to the lung^21^. CD14^+^ (CD16^+^/CD16^-^) cells have been shown to be capable of producing IL-8, IL-6, TNF-α, and IL-1 after activation, whereas CD14^dim^ CD16^+^ cells represent a monocyte subset that patrols blood vessels and selectively detects virally infected and damaged cells to produce proinflammatory cytokines^22^. Wei *et al*. recently reported the presence of inflammatory CD14^+^CD16^+^ monocytes, suggesting an excessive activated immune response caused by pathogenic GM-CSF^+^ Th1 cells, possibly linking them to the pulmonary immunopathology that leads to deleterious clinical manifestations after COVID-19 infections^23^.

Although it may be speculative to draw conclusions from serum cytokine profiles to interpret the pathophysiological phenomena of the lung, the large number of factors analyzed and their changes throughout the illness may provide crucial information. First, we found elevated levels of certain biomarkers already described as prognostic markers of severe disease. For example, elevated levels of IL-2, IL-7, IL-10, G-CSF, IP-10, MCP-1, MIP1-α, and TNF-α have been reported in SARS-CoV-2 patients in intensive care^3^. Increased Th17, cytotoxic markers^24^, IL-6 ^25^, IP-10, MCP-3, and IL-1RA ^26^ also appear to be associated with disease severity and fatal outcome in SARS-CoV-2-infected patients. Our data significantly extend these observations by the longitudinal survey of these changes. The various waves of markers suggest different stages, from a proinflammatory cytokine storm and Th1 and Th2 activation to T cell exhaustion, apoptosis, and cell cytotoxicity. It is difficult to correlate these features with clinical worsening from a single case, but a careful follow-up of these profiles in patients in ICUs may help to identify new prognostic markers or those of COVID-19 severity.

There is no currently validated antiviral treatment to control such severe SARS-CoV-2 infections. However, experimental drugs and drug combinations, such as Hydroxychloroquine, Remdesivir, Lopinavir-Ritonavir, or a combination of Lopinavir-Ritonavir and interferon-β -1b, are under investigation and may be considered for compassionate use for severely ill patients. Given the severity of the case, we tried various approaches, including an antiviral therapy and an immune therapy. Remdesivir did not accelerate the clearance of the virus from our patient. Coronaviruses are able to inhibit the interferon signaling pathways, which are, like those of T cell-mediated immunity, required for viral clearance^27^. Animal models and case reports suggest that the type-1 interferon-mediated response triggered by MERS-CoV may limit the viral disease to the lung and prevent systemic dissemination and viremia^28,29^. It is impossible to draw any conclusion on the effect of IFN-β in our case. Of note, we observed a dramatic decrease of IL-1RA from day 15 to day 22, contrasting with the persistence of high IL-1α and β expression. This discordant expression profile contrasts with preliminary observations in less critically ill patients (personal data). Although these observations were limited, the role of IL-1 as a pro-inflammatory cytokine and leukocyte migratory factor is very well known, suggesting the potential interest of IL-1RA in COVID-19 infection. Importantly, we found significantly higher expression of secreted IL-6, as have others, especially in ICU patients^23^, underscoring the rationale for testing anti-IL-6 or anti-IL-6R antibodies in severely-ill COVID-19 patients.

Our observations suffered from several serious limitations. First, we did not sample the patient at the very early phase of the disease. In addition, we only explored peripheral blood. Immune abnormalities of the other organs of the lymphoid compartments require further study. Nevertheless, we believe that this case report supports the worldwide effort to repurpose several marketed drugs and immunomodulators to reduce the inflammatory reaction and pathophysiology caused by the virus^30^. First, it clearly shows that the immune alterations are dynamic. In the first phase, the innate immune system, driven by monocytes, macrophages, and Tγδ lymphocytes, is activated and leads to T-lymphocyte exhaustion. The activation of B lymphocytes then contributes to a substantial humoral response. Finally, we observed endothelial activation. Overall, this individual case also underscores the need to combine two complementary approaches to limit the severity of the disease: to control viral replication with an antiviral, with the aim to limit the source of hyperinflammation, and to potentiate this effect using a specific immune modulator that targets the cytokine storm as a trigger of the pathophysiological response. These interventions should be evaluated in well-designed studies. This longitudinal analysis may help to determine the timing of such interventions and provide tools for the clinical follow-up of patients.

## Materials and Methods

### Patient and procedures

We report data for a patient who was admitted to the Bichat Claude Bernard ICU in January 2020. The investigation was conducted as part of an overall French clinical cohort assessing patients with COVID-19 and registered in clinicaltrials.gov under the following number: NCT04262921. It was approved by the French Ethics Committee and written informed consent was obtained from each patient involved. We used the open-source ‘Clinical Characterization Protocol for Severe Emerging Infections’ of the International Severe Acute Respiratory and Emerging Infection Consortium (ISARIC), supported by the World Health Organization (WHO), which has been updated in response to COVID-19.

We diagnosed SARS-CoV-2 by semi-quantitative reverse-transcriptase polymerase chain reaction (RT-PCR) on nasopharyngeal swabs in accordance with WHO guidelines.

Clinical, biological and radiological data were carefully recorded from computerized medical records as previously described. We also collected information on the dates of at-risk contact, the date of illness onset, and travel details in France.

### Virological data

We assessed the viral load from various samples (upper and lower respiratory tract, blood, urine and stool samples or rectal swabs, if appropriate, conjunctiva, and pleural effusion) according to the recommended protocols of ISARIC.

RNA extraction and real-time RT-PCR, primers and probes, high-throughput virus sequencing, virus isolation, and virus titration have been described elsewhere^10^.

### Cell phenotyping and quantification of serum analytes

PBMCs and serum samples were collected on days 14-16, 20, 22, and 24 after illness onset for evaluation of the various immunogenicity endpoints. All tests were performed at the Mondor Immunomonitoring Center, Vaccine Research Institute, Henri Mondor Hospital, Créteil, France.

Immune cell phenotyping was performed with an LSRII Fortessa 4-laser (488, 640, 561, and 405 nm) flow cytometer (BD Biosciences), and FlowJo software version 9.9.6 (Tree Star Inc.). CD4^+^ and CD8^+^ T cells were analyzed for CD45RA and CCR7 expression to identify the naive, memory, and effector cell subsets, for co-expression of activation (HLA-DR and CD38) and exhaustion/senescence (CD57and PD1) markers. CD19^+^ B-cell subsets were analyzed for the CD21 and CD27 markers. ASC (plasmablasts) were identified as CD19^+^ cells expressing CD38 and CD27. We used CD16, CD56, and Ki57 to identify NK-cell subsets. γδ T cells were identified using an anti-TCR γδ antibody.

A total of 72 analytes were quantified in heat-inactivated serum samples (see Fig. 3) by multiplex magnetic bead assays or ELISA. Serum from five healthy donors and five patients with septic shock sampled at D1 were also assayed as negative and positive controls, respectively. Two LXSAHM-2 kits, one for CD163 and ST2 and one for CD14 and LBP (R&D Systems); one LXSAHM-19 kit for IL-21, IL-23, IL-31, EGF, Flt-3 Ligand, Granzyme B, Granzyme A, IL-25, PD-L1/B7-H1, TGF-α, Aggrecan, 4-1BB/CD137, Fas, FasL,CCL-28, Chemerin, sCD40L, CXCL14, and Midkine (R&D Systems); and the 48-Plex Bio-Plex Pro Human Cytokines screening kit for IL-1β, IL-1rα, IL-2, IL-4, IL-5, IL-6, IL-7, IL-8 / CXCL8, IL-9, IL-10, IL-12 (p70), IL-13, IL-15, IL-17A / CTLA8, Basic FGF (FGF-2), Eotaxin / CCL11, G-CSF, GM-CSF, IFN-γ, IP-10/CXCL10, MCP-1 /CCL2, MIP-1α / CCL3, MIP-1β / CCL4, PDGF-BB (PDGF-AB/BB), RANTES/CCL5, TNF-α, VEFG (VEGF-A), IL-1a, IL-2Ra (IL-2R), IL-3, IL-12 (p40), IL-16, IL-18, CTACK / CCL27, GRO-a /CXCL1 (GRO), HGF, IFN-α2, LIF, MCP-3 / CCL7, M-CSF, MIF, MIG/CXCL9,b-NGF,SCF, SCGF-b,SDF-1α,TNF-b/LTA, and TRAIL (BioRad) were used according to the manufacturers’ recommendations and data were acquired on a Bio-Plex 200 system™. FABP2/IFABP quantification was performed with the Human Quantikine ELISA Kit (R&D Systems), according to the manufacturers’ instructions. Extrapolated concentration values were considered and the out of range values were entered at the highest or lowest extrapolated concentration. Expression values were standardized for each cytokine for all displayed samples (centered around the observed mean, with variance equal to 1). Hierarchical clustering of the cytokines was computed using the Euclidean distance and Ward’s method^31^ for all displayed samples.

## Data Availability

All the datasets that support the findings of this study are available from the corresponding author on reasonable request.

## Acknowledgements

We thank French COVID 19 cohort for the cohort management and sample collection

## Author Contributions

LB, JFT and YL conceived and designed the study. LB, JP, PHW, JLD, FS, EdM and JFT participated in sample collection and clinical follow-up of the patient. EF, MS and AW performed the experiments. YL, JFT, LB, AW, RT, MS, BH, CL analyzed and interpreted the data. All authors approved the final version.

